# The Most Cost-Efficient And Effective Set Of Covid-19 Policies To Reduce Monthly Covid-19 Case Increase Rates

**DOI:** 10.1101/2022.09.04.22279016

**Authors:** Charline Chen

## Abstract

The coronavirus is one of the most unprecedented pandemics in recent decades. Countries have been struggling to identify the appropriate policies to prevent COVID-19 spread efficiently. As coronavirus case and death numbers fluctuated among countries in the past two years, questions of which policies are most cost-efficient and effective in preventing coronavirus spread have yet to be answered. There are no worldwide agreed guidelines to follow, to the author’s best knowledge. Countries are prone to making policy and implementation errors that could cost lives and cause tremendous economic loss. Although much research on COVID-19 has been done and many focused on policy effectiveness, few focused on the cost-efficiency and effectiveness of COVID-19 policies. This research identifies the most Cost-Efficient and Effective Set of COVID-19 Policies to Reduce Monthly COVID-19 Case Increase Rates through a quantitative, big-data-driven, and machine-learning enabled approach. The research collected and analyzed 13 COVID-19 policies and associated daily COVID-19 case numbers across 180 countries from January 2020 to June 2021, developed Policy Cost Model, defined Policy Efficiency and Effectiveness Index, and developed a Fully-Automated Best Policy Group Finder Python Program to find the most cost efficient and effective policy group. This research found that 1) Before Vaccinations are available, the most cost-efficient and effective policy group includes Facial Covering, Testing Policy, and Contact Tracing. Its cost-efficiency is a 1% monthly case decrease rate per billion of dollars spent. Its effectiveness is a 37% monthly case decrease rate. It is 1474 times more cost-efficient, 11 times more effective, and costs around $5336.83 Billion less than implementing all 12 common COVID-19 policies as the U.S. and many other countries did before vaccinations were available; 2) After Vaccinations are available, the most cost-efficient and effective policy group includes Facial Coverings, Contact Tracing, and Vaccinations. Its cost-efficiency is a 2.7% monthly case decrease rate per billion of dollars spent. Its effectiveness is a 52% monthly case decrease rate. It is 3835 times more cost-efficient, 21.5 times more effective, and costs around $5350 Billion less than implementing all 13 common COVID-19 policies as the U.S. and many other countries did. The research results will help countries, especially underdeveloped ones with very limited budgets, to identify and implement the most cost-efficient and effective policies so they can spend much less but reduce monthly cases much more.

## 1. Introduction

### 1.1 Background

COVID-19 was first discovered in Wuhan, China, in December 2019. Since then, there have been around six million deaths and 528 million cases of COVID-19 worldwide as of May 2022. Countries worldwide have spent trillions of dollars to fight the pandemic. Although COVID-19 has had tremendous impacts on people’s lives and economies, there is still no worldwide agreement on the most cost-efficient and effective policies to implement, and guidelines for countries to follow. There are countries like Sweden whose government never ordered a full lockdown [20], countries like China who have a very strict zero-COVID policy, and a majority of countries that follow policies whose strictness are somewhere in between. For example, the U.S. enforced some COVID-19 policies (e.g. face-mask, school shutdowns, travel restrictions, etc.) to a certain degree in the past two years, and began to relax them in 2022. While COVID-19 death counts are still rising and the global economy is still suffering from a broken supply chain and inflation(partially caused by this pandemic), countries and politicians are still debating on what the most cost-efficient and effective policies to combat COVID-19 are. In addition, most developed countries spend as much as they want to fight the pandemic without trying to identify the most efficient and effective policies, hence wasting a lot of money. On the other hand, underdeveloped countries with very limited budgets picked policies to implement through only educated-guesses; they did not know what the most cost efficient and effective policy group was. As a result, more deaths, suffering, and economic costs occurred. This research utilized big data analysis and a machine-learning enabled approach to identify the most cost-efficient and effective COVID-19 policy group so that countries can quickly identify and implement them to minimize the pandemic’s economic impact and save more lives.

### 1.2 Problem

Much research has been done to identify the most effective COVID-19 policies or predict COVID-19 cases through utilizing machine learning algorithms like Bayesian analysis or Poisson regression. However, the scope of these researches is quite limited, such as focusing only on the U.S., which is not representative of the worldwide situation. Furthermore, most do not consider the cost of each policy, which is a critical success factor in a country’s practice of combating COVID-19, especially for underdeveloped countries with very limited budgets. Therefore, those researches are not able to identify which COVID-19 policies are the most cost-efficient and effective across the world, and their results are not sufficient to be established as a common worldwide guideline for countries to follow.

### 1.3 Solution

To identify the most cost-efficient and effective COVID-19 policy group in reducing monthly case increase rates, and ensure it is applicable for countries worldwide, my research: 1) collected data from ourworldindata.org, which has data across 180 countries on 13 common policies and COVID-19 cases numbers for around two years; 2) defined a policy cost model, cost-efficiency model, effectiveness model, and a cost-efficiency and effectiveness index to measure a policy group’s integrated efficiency and effectiveness; and 3) developed a machine learning based program to loop through all possible combinations of the 13 common COVID-19 policies to calculate and identify the most cost-efficient policy group, the most effective policy group, and the most cost-efficient and effective policy group. Because this research is based on worldwide data(across 180 countries) collected from ourworldindata.org, the data-driven analysis conclusions should provide meaningful policy guidelines in combating COVID-19 to countries around the world. Further, because the research provided a well-defined policy cost model, cost-efficiency model, and effectiveness model, and programmatically calculated policy groups’ efficiency and effectiveness across all 8191 possible combinations of 13 common policies to identify the most cost-efficient and effective policy group, the research result should provide practical guidelines to countries worldwide in implementing the most cost-efficient and effective policy group to reduce COVID-19 monthly case increase rate.

### 1.4 Experiment

To identify the most cost-efficient and effective policy group in a quantitative way, the research first defined a cost model for each policy, and the calculation formulas for measuring every policy group’s cost-efficiency, effectiveness, and integrated cost-efficiency and effectiveness index. Then, a machine-learning Python program was developed and run to automatically identify a best linear regression monthly case increase rate prediction model; the prediction model function was then used to calculate and find the most cost-efficient policy group, the most effective policy group, and the most cost-efficient and effective policy group respectively from all 8191 possible combinations of the 13 policies.

The accuracy of the monthly case increase rate machine learning model was evaluated using R-Squared, which is a representation of how well the model fits the dataset. The R-Squared of the group of policies had a range of around 0.03 to 0.19. Though they may seem relatively low, it is actually not bad since this research is highly related to an observation of human behavior. Since human behavior is very difficult to be exactly predicted, machine learning models regarding human behavior usually have lower R-Squared values.

The research ran the program in six batches - 5 times (once), 10 times (twice), and 50 times (three times) - to identify the most common best possible groups of policies. The results are quite consistent. Among the six batch runs, five of them identified the same best policy groups for the most cost-efficient group, and the most cost-efficient and effective policy group.

### 1.5 Paper Structure

The rest of the paper is organized as follows: Section 2 discusses the challenges I encountered during research and experimentation; Section 3 focuses on the details of the solutions corresponding to the challenges mentioned in Section 2; Section 4 presents the relevant details about the experiments done. Section 5 briefly discusses some related research, the difference between those researches and mine, and identifies key contributions this research will add. Finally, Section 6 gives concluding remarks, as well as discussing future work.

## 2. Challenges

### 2.1 Challenge 1: Policy Cost Model

One of the greatest challenges in this project was to identify and develop a policy’s cost model. Because there is currently no comprehensive and solid government-released data on the cost of each of the common 13 COVID-19 policies(facial covering, testing policy, contact tracing, vaccination, internal movement restrictions, international travel controls, public information campaigns, public event cancellations, gathering restrictions, closing public transportation, school closures, stay at home requirements, workplace closures), there is very little information available about costs for any of the 13 common COVID-19 policies. This is also likely the reason that most COVID-19 policy research has focused on policy effectiveness or case predictions, while little research has been done on policy cost-efficiency.

### 2.2 Challenge 2: Diverse and Inclusive Research Results for Underdeveloped Countries

To ensure the research’s result is appropriate to provide meaningful guidelines worldwide, the research needs good-quality worldwide data across countries. However, although most developed countries have shared consistent higher-quality data in COVID-19 policies and case numbers, many underdeveloped countries have only provided COVID-19 data for limited periods and quite often with some missing data. That is likely the reason that quite some research focused only on developed countries based on the large amounts of data collected from them. It is critical for the research to collect and identify COVID-19 policies and case numbers from a wide variety of countries worldwide - across developed and underdeveloped ones - in a common time period so that the research can provide practical worldwide guidelines.

### 2.3 Challenge 3: Most Cost-Efficient AND Effective policy group

Quite often, the most effective policy group for COVID-19 monthly case decrease rate is not the same as the most cost-efficient policy group. The third challenge was to identify the most cost-efficient and effective policy group in a consistent and systematic way. In the real world, few countries can implement COVID-19 policies by only focusing on the most effective policy group without considering the cost. Hence, it is critical for the research to identify the most cost-efficient and effective policy group so that it can provide pragmatic guidelines for countries, especially underdeveloped ones, to identify and implement the most cost-efficient and effective policies within their limited budget so that they can spend less while reducing monthly COVID-19 increase rates more.

## 3. Methodology/Solution

### 3.1 Overview of the Solution

To answer the research question(Which group of COVID-19 policies are the most cost-efficient and effective to reduce monthly case increase rates?), the research set the hypothesis: If we implement all policies negatively correlated to the monthly case increase rate, then we will achieve the highest effectiveness of reducing the monthly COVID-19 case increase rate with the greatest efficiency. The research took the following approach (as shown in Figure 1): 1) Research and identify raw data sources including COVID-19 policies and monthly case numbers in all available countries (around 180); 2) Data Engineering to clean and normalize the data to identify consistent and qualified COVID-19 policies and case numbers data across 169 countries in 15 months. This will address challenge #2 (Diverse and Inclusive Research) listed in section 2; 3) Data Analysis to identify each policy’s correlation with monthly case increase rate; 4) Research, Develop and Define Policy Cost Model, Cost Efficiency Model, Effectiveness Model, and Most Cost-Efficient and Effective policy group Index. This will address challenge #1 (Policy Cost Model) and #3 (Most Cost Efficient AND Effective policy group) challenges listed in section 2; 5) Develop Fully-Automated Best Policy Group Finder Python Program, and Run it to Find the Most Cost-Efficient, Most Effective, and Most Cost-Efficient and Effective policy group(s). This will further address challenge #3 listed in section 2; 6) Analyze major policy groups(including most efficient, most effective, and most efficient AND effective group) to identify practical recommendations.

**Figure 1.**
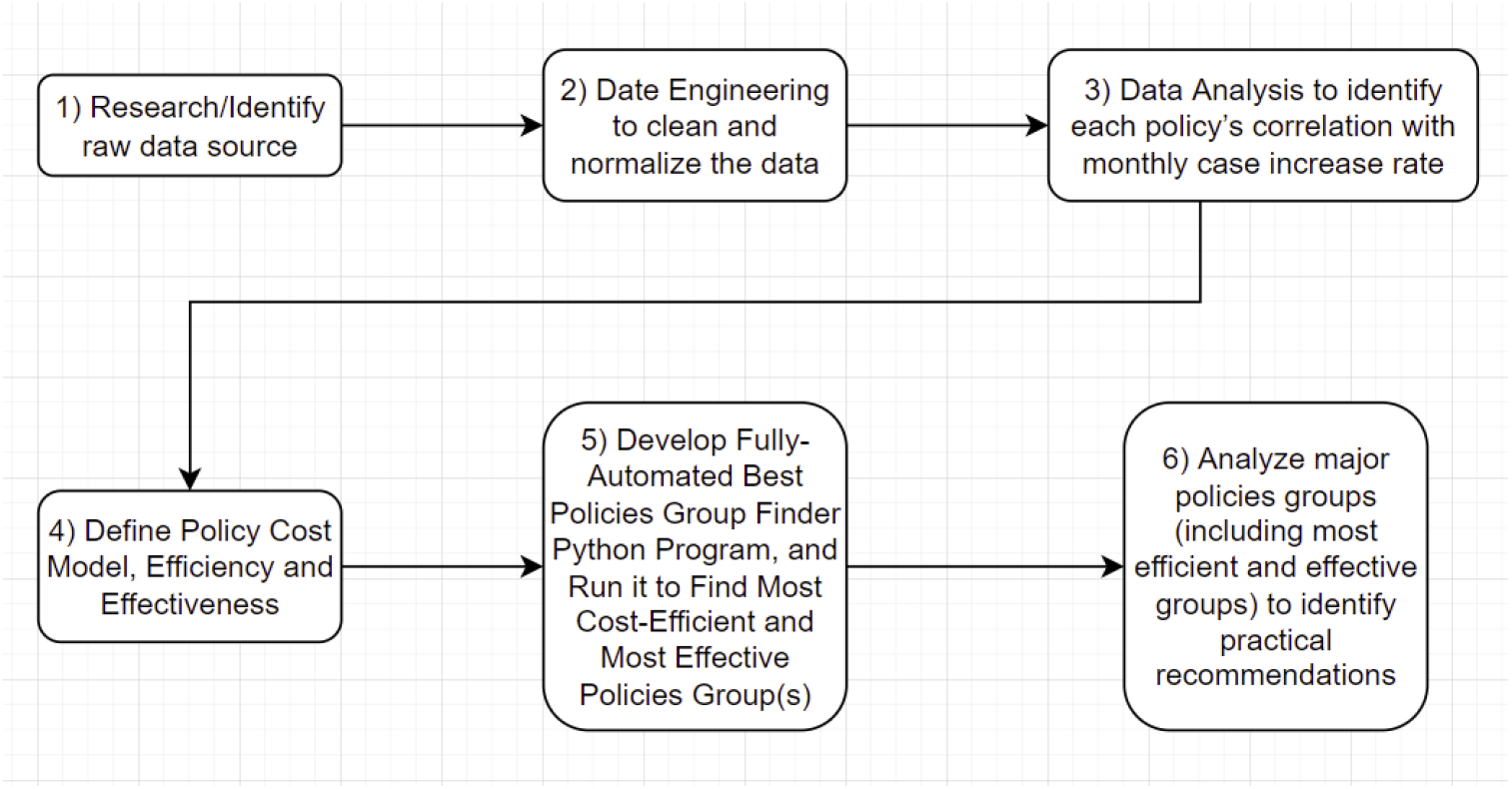
Research Approach Overview

### 3.2 Technical Details by Modules

This research utilized Python Numpy’s correlation function to first identify the policies that were negatively correlated with the monthly case increase rate. These policies were grouped as the All-Negative-Correlated Polices Group, which includes: Facial Coverings, Testing Policy, Vaccination Policy, Contact Tracing, Public Information Campaigns, and Gathering Restrictions. The hypothesis was set as: If we implement all policies negatively correlated to the monthly case increase rate, then we will achieve the highest effectiveness of reducing the monthly COVID-19 case increase rate with the greatest cost efficiency.

Then, to identify the most cost-efficient and effective COVID-19 policy group in a quantitative and systematic way, the research took the following steps.

First, the research collected the raw data(from ourworldindata.org) on the 13 policies’ enforcement levels and daily COVID-19 case numbers in 180 countries from January 2020 to June 2021. This helped ensure the data in the research would not be biased toward advanced countries. Further, the research analyzed all the data and noticed that some countries’ policy and case numbers had null values, or were lacking data in some months. This could be because some underdeveloped countries did not have the resources to collect their COVID-19 policy and case numbers, or that some countries had not even begun to monitor COVID-19 in early 2020. Through further analysis of the data collected, the research identified around 169 countries having consistent COVID-19 policy and case numbers in the period from April 2020 - May 2021 because it was a period of time when the majority of countries had already been impacted by the pandemic and rushed to monitor COVID-19.

Second, after the raw data was uploaded to Google Colab, a Python program was developed to clean, process, and normalize the data. To utilize Linear Regression Algorithm, the data was normalized by a Python program to calculate the average monthly case number, average monthly policy implementation enforcement level, and monthly case increase rate. In the dataset, a column called “Monthly Case Increase Rate” was added. The formula to calculate it is (Month 2’s Total Cases - Month 1’s Total Cases) / Month 1’s Total Cases. The resulting increase rate would be placed as Month 1’s Monthly Case Increase Rate. However, there was a problem when the data went from the last month(e.g. May 2021) of a country, to the first month(e.g. April 2020) of the next country. There was no data for June 2021 that could be used to identify May 2021’s monthly case increase rate. Thus, this research sets May 2021’s Monthly Case Increase Rate as April 2021’s Monthly Case Increase Rate. Although this increase rate calculated for the last month (e.g. May 2021) is an approximation, it should have very limited impact on the overall machine learning model and data analysis because it only accounts for less than 8% of total data, and the approximation should be close to the actual data.

Third, another Python program was developed to identify each policy’s correlation with the monthly case increase rate, and create a correlation heat map/matrix for each policy against all other policies, as Figure 2 and 3 show below.

**Figure 2:**
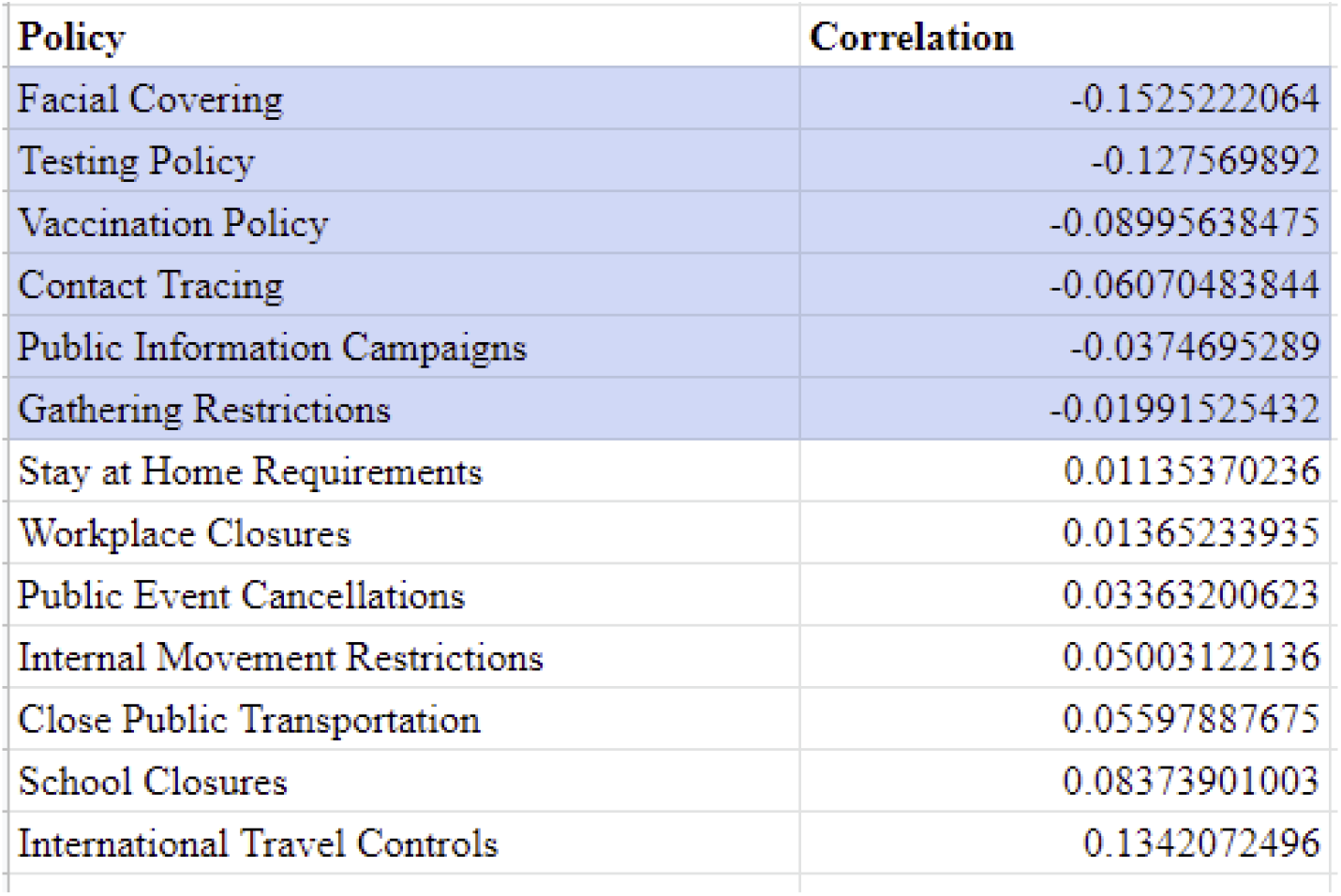
Correlation Table of Policies against Monthly Case Increase Rate

**Figure 3:**
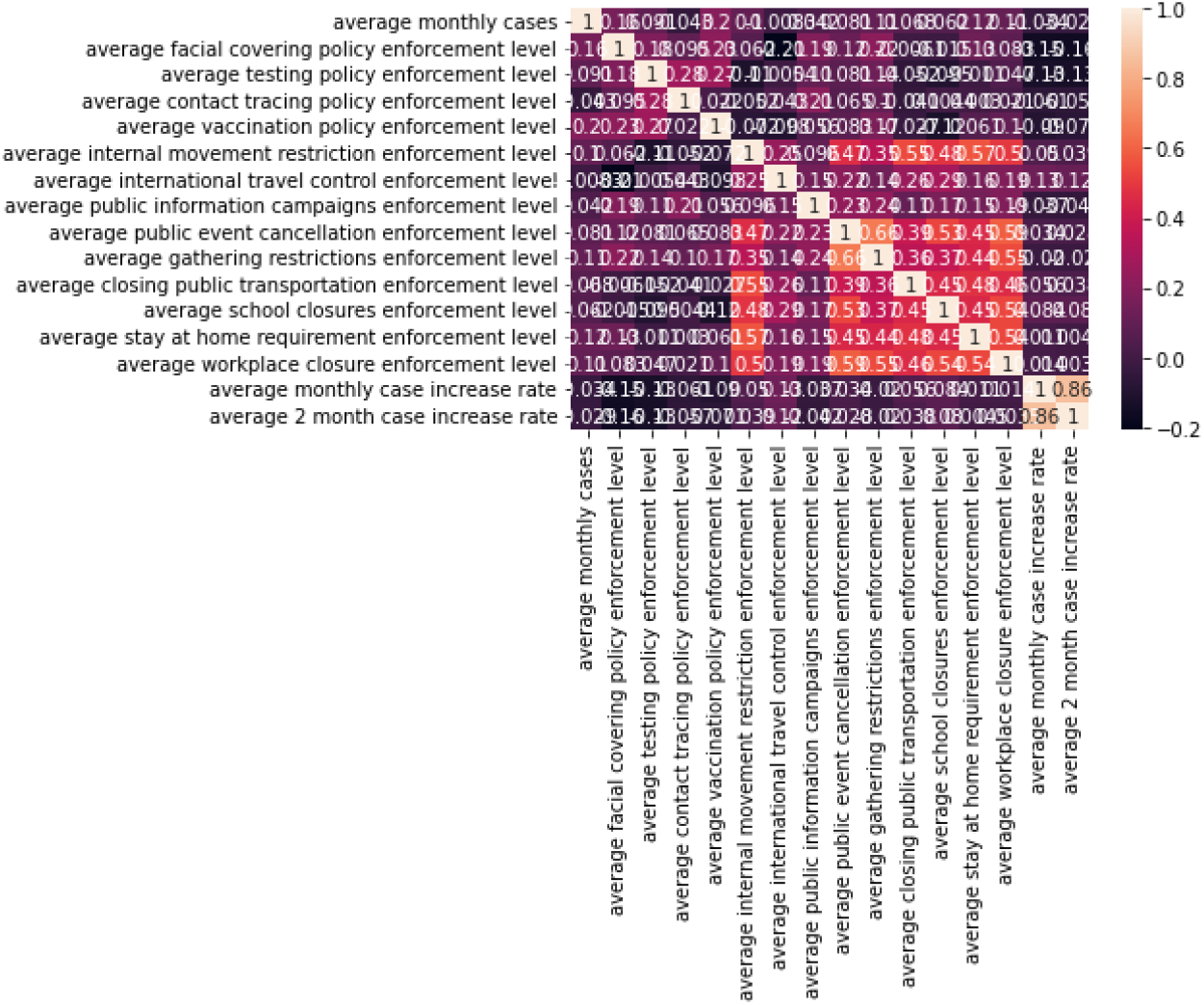
Correlation Heatmap of Policies

Fourth, to find the most cost-efficient and effective policy group, the research defined the following:

1. The cost model[2, 5-12, 14-19], as shown in Figure 4, is an estimation of the annual cost of each of the 13 policies based on the cost data that could be found and estimated in the U.S. Extensive search, analysis, and estimation on the annual cost of each of the 13 policies in the U.S. has been done. The research utilized many different sources and logical reasoning to estimate each policy’s annual cost. For example, for the facial covering policy, research on Amazon found that the cost of 50 face masks = $4.39 + tax ∼=$5.00. Then, looking at the U.S. population being around 333, 315, 463(on the day of the cost model creation) and assuming that every citizen uses one mask per day, the total cost of the facial coverings policy would be: U.S. population * unit price of face mask * 365 days ∼= 12.17 billion. Some policy costs are more difficult to estimate, like those for the Internal Movement Restrictions, Public Event Cancellations, and Gathering Restrictions policies. For these three, because they are all related to the costs of restaurants closures, International Travel Restrictions, and Public Transportation Restrictions, the research collected available cost data on Restaurants Closures, International Travel Restrictions, and Public Transportation Restrictions, summed them up, and divided the total by three. Thus, an estimated average annual cost of $137 billion was established for each of the three policies - Internal Movement Restrictions, Public Event Cancellations, and Gathering Restrictions.
2. Effectiveness of a policy group (measured by the decrease rate, or negative increase rate, of monthly cases) = (Average Daily Case Number in Month 2 - Average Daily Case Number in Month 1) / (Average Daily Case Number in Month 1)
3. Efficiency of a policy group (measured by the decrease rate of monthly cases per billion of dollars) = Effectiveness of the policy group / Total Annual Cost of the Policy Group, where Total Annual Cost of the Policy Group = The Sum of the Cost of Each Policy in the Policy Group
4. Integrated Cost-Efficiency and Effective Index = [((Cost-Efficiency * -1) / Maximum Cost Efficiency) * Efficiency-Weight] + [(Effectiveness * -1) * Effectiveness-Weight], where Maximum Cost Efficiency = 4%/$Billion, Efficiency-Weight = 0.4 and Effectiveness-Weight = 0.6, so that the research can identify the most cost-efficient and effective policy group in a well-defined quantitative way
  a. The *-1 is included because the cost-efficiency and effectiveness values are usually negative since they represent a decrease in monthly COVID-19 cases. But for the sake of our index value, we will want a positive value.
  b. The Maximum Cost-Efficiency is estimated to be “0.04” based on hundreds of program runs. Cost-Efficiency divided by Maximum Cost-Efficiency is used to normalize/scale a policy group’s cost efficiency appropriately to align with the scale of effectiveness (0-100%). There is no need to rescale a policy group’s Effectiveness since the maximum possible effectiveness is 100% (a decrease rate of 100%) and the effectiveness values are already calculated at that scale.
  c. The cost-efficiency weight is 0.4 while the effectiveness weight is 0.6 because countries would usually focus more on effectiveness over cost-efficiency.

**Figure 4:**
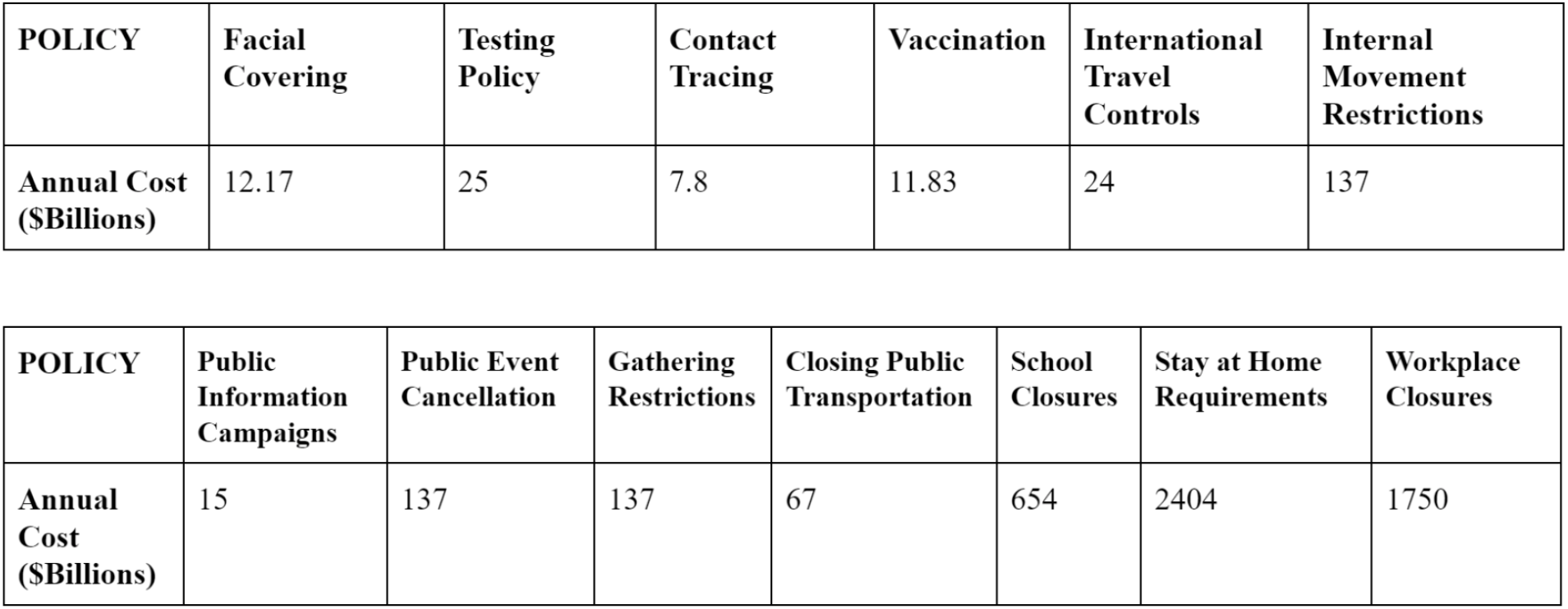
Policy Cost Model

Fifth, the third Python program is developed and run to automatically find the best combination of policies for the Best Prediction Linear Regression Machine Learning Model of Monthly Increase Rate, Most Effective policy group, Most Cost-Efficient policy group, and Most Cost-Efficient AND Effective policy group respectively from all 8191 possible combinations of the 13 policies.

As outlined in Figure 5, the third Python program algorithm flow is: 1) The first loop will loop through 1, 2, 3, …, 13 number of policies 1.1) Under the first loop, calculate the number of combinations for the number of policies 1.2) The second (inner) loop will loop through the number of combinations in the current number of policies in the first loop 1.2.1) Through utilizing Google Colab, Python, sklearn library, and integrated policy related dataset, the research created a machine learning model, calculated R-Squared of the model, the effectiveness, the cost-efficiency, and integrated efficiency and effectiveness index for the current combination of policies. The dataset including the Vaccination Policy was split into the X dataset with predictor variables(the policies’ enforcement levels) and the y dataset with the prediction variable(the monthly case increase rate). The two datasets were split into training and testing datasets with an 80%-20% split. The training X and y datasets were used to develop the Linear

**Figure 5:**
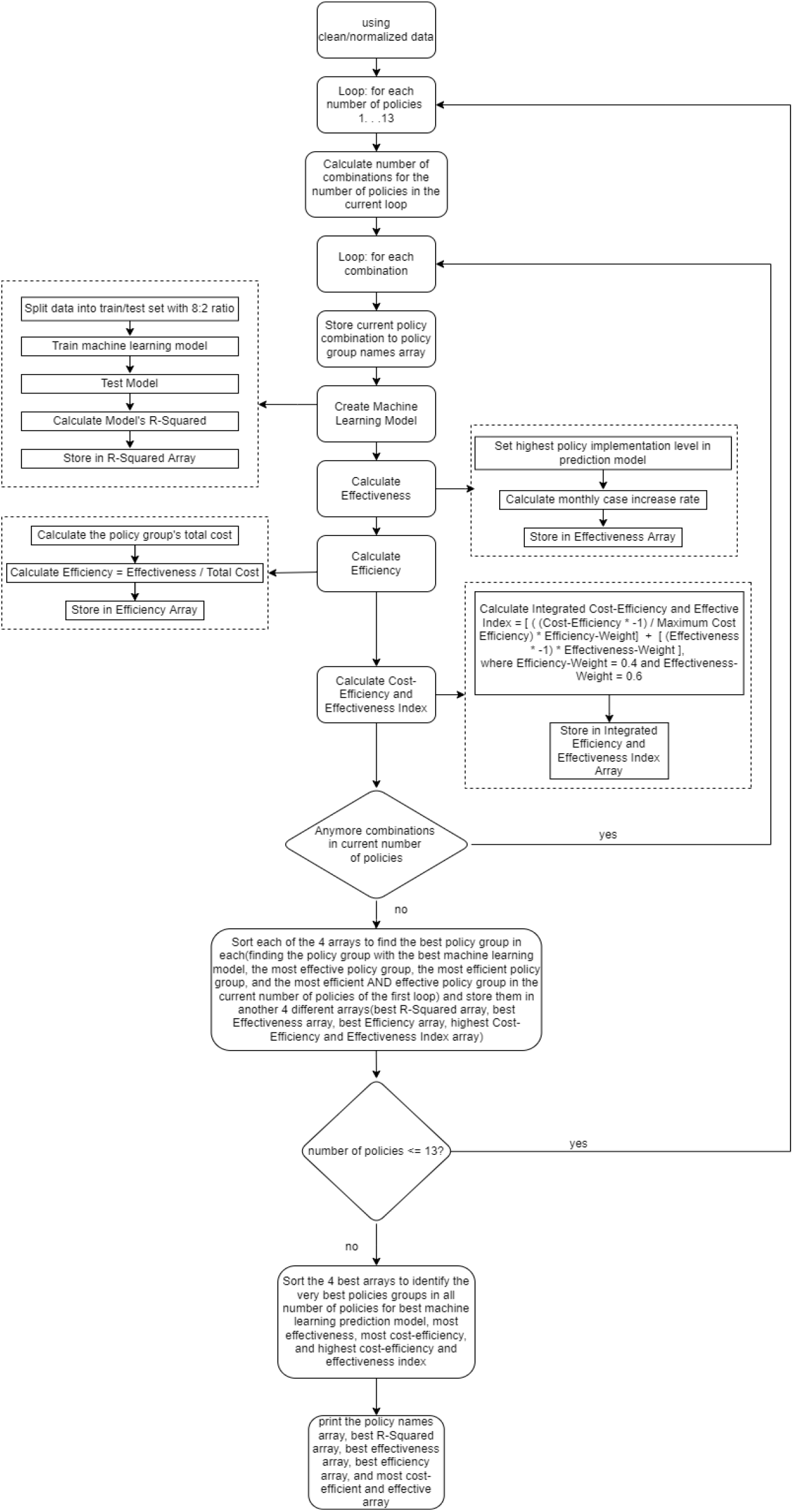
Best policy group Finder Algorithm Flow

Regression model for monthly case increase rate predictions. The Linear Regression model was then used to find the cost-efficiency and effectiveness of a policy group. 1.2.2) Store the R-Squared, effectiveness, efficiency, and integrated efficiency and effectiveness index of the current combination of policies in a different array respectively 1.3) Sort the four arrays defined in 1.2.2 to find the best policy group in each of the four arrays to identify the best machine learning prediction model, most effectiveness, most cost-efficiency, and most efficient and effective ones in the current number of policies of the first loop, and store them in another four different arrays 2) After the loops, sort the four arrays defined in 1.3 to identify the best policy groups in all number of policies combinations for the best machine learning prediction model, most effective policy group, most cost-efficient policy group, and most cost-efficient and effective policy group.

Sixth, the relationships between major policy groups (the most cost-efficient group, most effective group, most cost-efficient and effective group, all negative correlated policy group, and all 13-policy group) are analyzed(in section 4.3), and pragmatic, meaningful COVID-19 policies recommendations are concluded (in section 6)

## 4. Experiments/Evaluation

### 4.1 Finding the Best Policy Groups Among 13 Policies (Including Vaccinations)

#### 4.1.1 Setup

The first experiment is based on the 13 policies data including the Vaccination policy to find the most cost-efficient policy group, most effective policy group, and most cost-efficient and effective policy group.

#### 4.1.2 Results

The Best Policies Finder program was run in six batches: 5 times(once), 10 times(twice), and 50 times(three times). Five of the six batches found the same policy groups for most efficient, and most efficient and effective. The following table shows the results.

As the result table shows:

- The most cost-efficient policy group consisted of Contact Tracing and Vaccination Policy. It has a cost-efficiency of around a 2.7% monthly case decrease rate per billion of dollars spent, an effectiveness of around 52%, and a cost-efficiency and effectiveness index of around 0.58.
- The most effective policy group consisted of Facial Coverings, Testing Policy, Contact Tracing, Vaccinations, Public Information Campaigns, Public Event Cancellations, and Stay at Home Requirements. It has an effectiveness of around 86%, a cost-efficiency of around a 0.03% monthly case decrease rate per billion of dollars spent, and a cost-efficiency and effectiveness index of around 0.52.
- The most cost efficient and effective policy group consists of Facial Coverings, Contact Tracing, and Vaccination. It has a cost-efficiency of around 2.3%, an effectiveness of around 74%, and Cost-Efficiency and Effectiveness Index of around 0.68. Compared to the most cost-efficient policy group, this group with the inclusion of Facial Coverings had a cost-efficiency that remained similar (2.3%/$B) as the most cost efficient group’s (2.7%/$B), but the effectiveness greatly increased from 52% to 74%.

### 4.2 Finding the Best Policy Groups Among 12 Policies (Without Vaccinations)

#### 4.2.1 Setup

The second experiment is based on 12 policies’ data excluding vaccine policy to find the most cost-efficient policy group, most effective policy group, and most cost-efficient and effective policy group. Because vaccines will not be available when most pandemics start, it would be meaningful to analyze data and identify the most cost-efficient and effective policies when vaccines are not yet available. The Best policy group Finder Algorithm applied in this experiment is the same one as the first experiment.

#### 4.2.2 Results

The Best Policies Finder program was run in six batches: 5 times(once), 10 times(twice), and 50 times(three times). Five of the six batches find the same policy groups for most efficient, and most efficient and effective. The following table shows the results:

As the result table shows:

- The most cost-efficient policy group consisted of Facial Coverings and Testing Policy. It has a cost-efficiency of around a 1% monthly case decrease rate per billion of dollars spent, an effectiveness of around 37%, and a cost-efficiency and effectiveness index of around 0.32.
- The most effective policy group consisted of Facial Coverings, Testing Policy, Contact Tracing, Vaccinations, Public Event Cancellations, and Stay at Home Requirements. It has an effectiveness of around 42%, a cost-efficiency of around a 0.02% monthly case decrease rate per billion of dollars spent, and a cost-efficiency and effectiveness index of around 0.25.
- The most cost efficient and effective policy group consists of Facial Coverings, Testing Policy, Contact Tracing. It has a cost-efficiency of around 0.9%, an effectiveness of around 40%, and Cost-Efficiency and Effectiveness Index of around 0.33.

### 4.3 Analysis

Both experiments ran in six batches. Five of the six batch-runs in each experiment identified the same policy group for most efficient, and most efficient and effective one, respectively.

As shown in Table 3, this research further analyzed and compared the most cost-efficient, most effective, and most cost-efficient and effective policy group with the All-Negative-Correlated policy group and the All-13 policy group. The All-Negative-Correlated policy group’s cost-efficiency is around a 0.3% monthly case decrease rate per billion dollars spent, and has an effectiveness of around a 65% monthly case decrease rate. When looking at all 13 COVID-19 policies, the cost-efficiency is around 0.00061% while the effectiveness is around 3.3%. These two policy groups have much lower cost-efficiency and effectiveness values compared to the most cost-efficient and effective policy group. The Negative Correlation Policy Group costs around $177 billion more, and the All 13 Policy Group costs around $5350 billion more than the most cost-efficient and effective policy group identified in Experiment 1.

**Table 1:**

|  |  |
| --- | --- |
| **Most Cost-Efficient** | **RSQ:** -0.1062300884715337<br>**Policies:** [['average contact tracing policy enforcement level'], ['average vaccination policy enforcement level']]<br>**Coefficients:** [array([[ -0.40856777, -0.27319009]]), array([1.6619468])]<br>**Effectiveness:** [-0.52113922]<br>**Efficiency:** [-0.0265481] |
| | **Total Cost:** \$19.63 Billion<br>**Effectiveness and Efficiency Index:** [0.57816454] |
| **Most Effective** | **RSQ:** -0.10710021129695879<br>**Policies:** [['average facial covering policy enforcement level'], ['average testing policy enforcement level'], ['average contact tracing policy enforcement level'], ['average vaccination policy enforcement level'], ['average public information campaigns enforcement level'], ['average public event cancellation enforcement level'], ['average stay at home requirement enforcement level']]<br>**Coefficients:** [array([[ -0.47083448, -0.42594843, -0.20106711, -0.12456212, -0.15991866, 0.39701597, -0.07153428]]), array([3.06585341])]<br>**Effectiveness:** [-0.86068284]<br>**Efficiency:** [-0.00032941]<br>**Total Cost:** \$2612.8 Billion<br>**Effectiveness and Efficiency Index:** [0.5197038] |
| **Best Machine Learning Model** | **RSQ:** 0.18138002544666643<br>**Policies:** [['average facial covering policy enforcement level'], ['average testing policy enforcement level'], ['average contact tracing policy enforcement level'], ['average vaccination policy enforcement level'], ['average internal movement restriction enforcement level'], ['average international travel control enforcement level'], ['average closing public transportation enforcement level'], ['average school closures enforcement level']]<br>**Coefficients:** [array([[ -0.29668132, -0.4236867 , -0.19571087, -0.08340682, -0.07343047, 0.3462473 , 0.01925134, 0.12498792]]), array([1.61891428])] |
| **Largest Integrated Cost-Efficiency and Effectiveness Index** | **RSQ:** -0.2897973254484161<br>**Policies:** [['average facial covering policy enforcement level'], ['average contact tracing policy enforcement level'], ['average vaccination policy enforcement level']]<br>**Coefficients:** [array([[ -0.48551165, -0.34197414, -0.16901528]]), array([2.72673874])]<br>**Effectiveness:** [-0.74433255]<br>**Efficiency:** [-0.02340668]<br>**Total Cost:** \$31.8 Billion<br>**Effectiveness and Efficiency Index:** [0.68066637] |

**Table 2:**
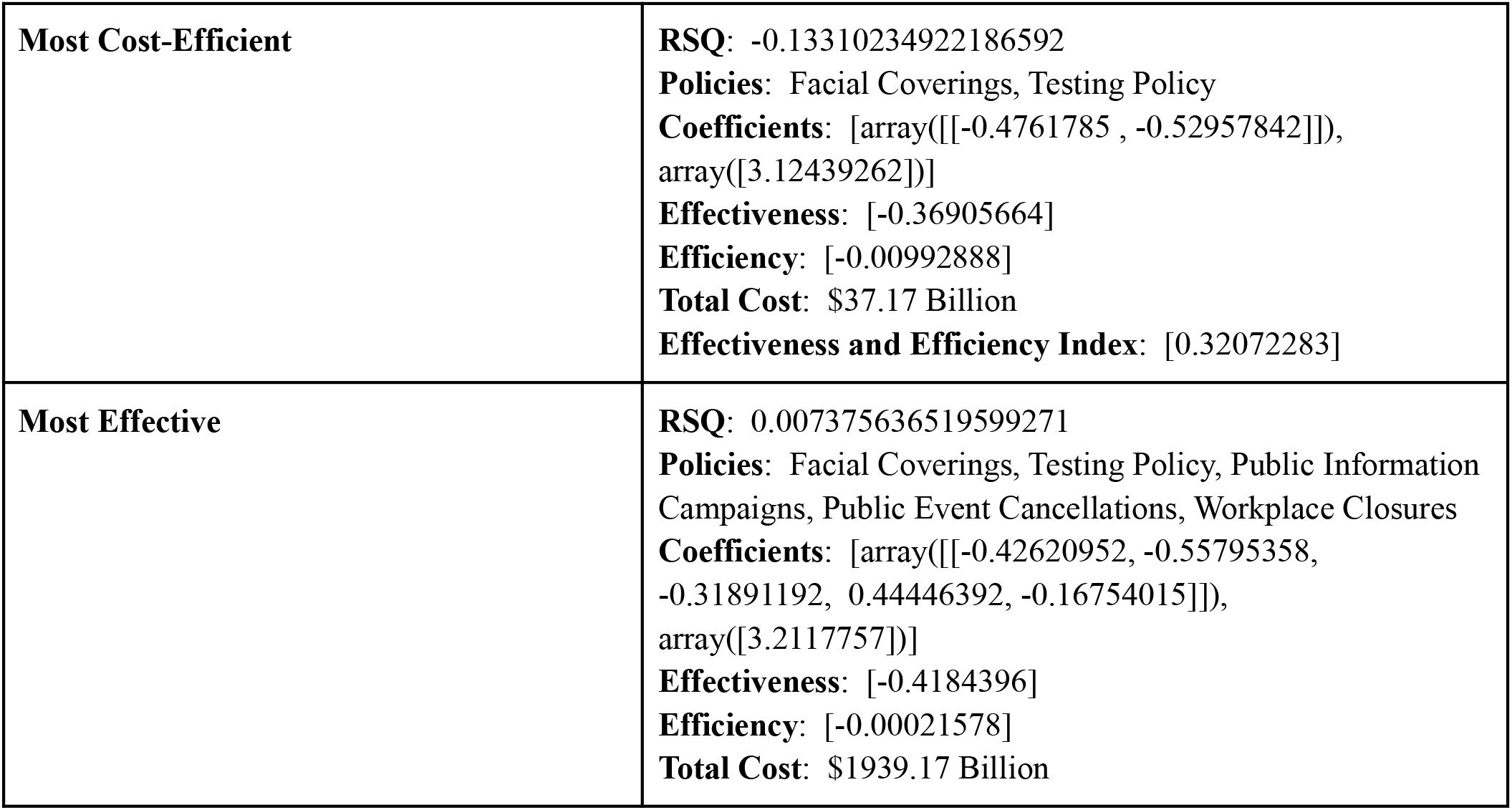
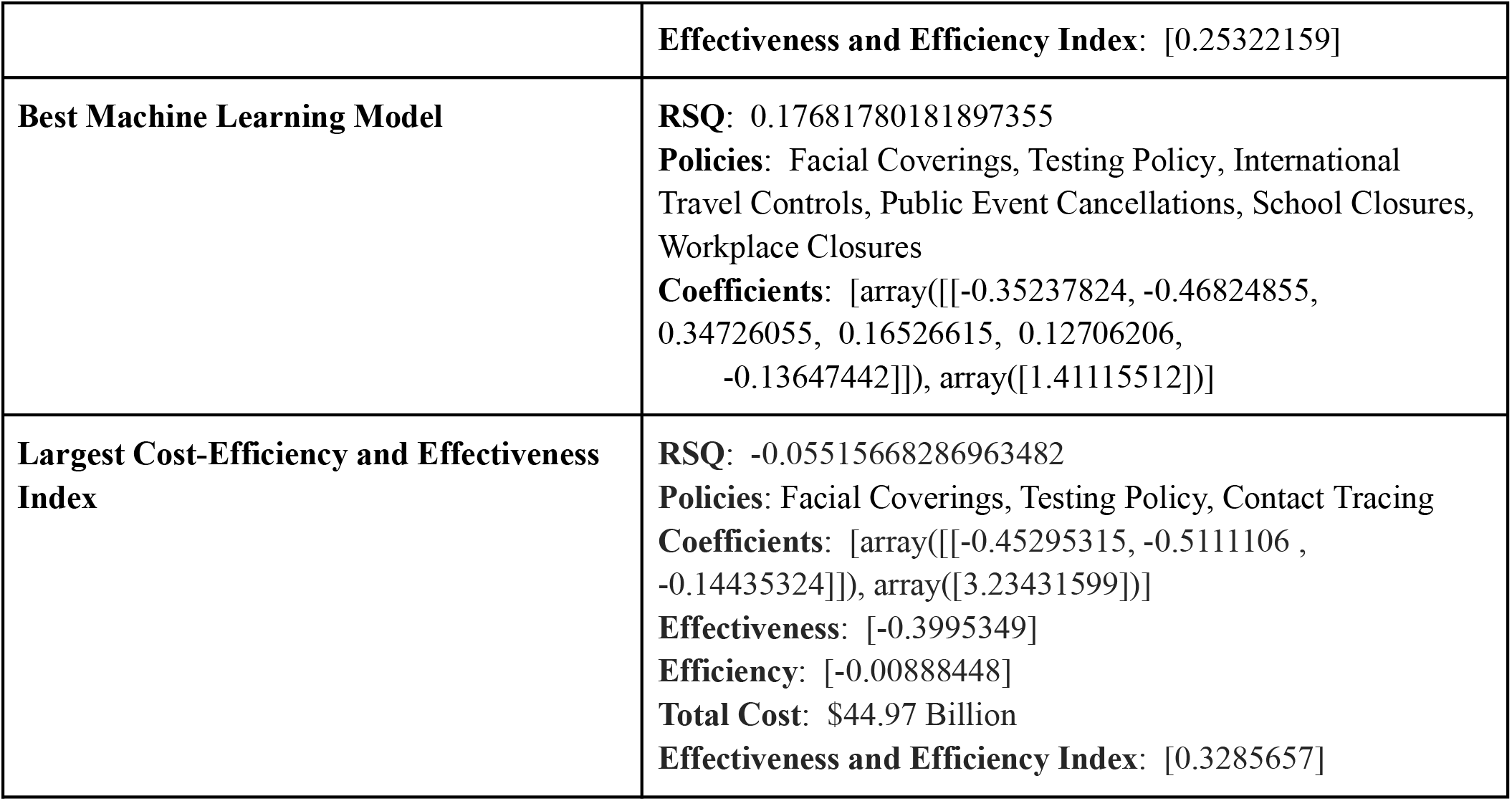

**Table 3:** Five Major Policy Groups Comparison Table

|  |  |
| --- | --- |
| **Most Cost-Efficient policy group** | **Policies:** [['average contact tracing policy enforcement level'], ['average vaccination policy enforcement level']]<br>**Effectiveness:** [-0.52113922]<br>**Efficiency:** [-0.0265481]<br>**Total Cost:** \$19.63 Billion<br>**Effectiveness and Efficiency Index:** [0.57816454] |
| **Most Effective policy group** | **Policies:** [['average facial covering policy enforcement level'], ['average testing policy enforcement level'], ['average contact tracing policy enforcement level'], ['average vaccination policy enforcement level'], ['average public information campaigns enforcement level'], ['average public event cancellation enforcement level'], ['average stay at home requirement enforcement level']]<br>**Effectiveness:** [-0.86068284]<br>**Efficiency:** [-0.00032941]<br>**Total Cost:** \$2612.8 Billion<br>**Effectiveness and Efficiency Index:** [0.5197038] |
| **Most Cost-Efficient AND Effective policy group** | **Policies:** [['average facial covering policy enforcement level'], ['average contact tracing policy enforcement level'], ['average vaccination policy enforcement level']]<br>**Effectiveness:** [-0.74433255]<br>**Efficiency:** [-0.02340668]<br>**Total Cost:** \$31.8 Billion<br>**Effectiveness and Efficiency Index:** [0.68066637] |
| **All Negative Correlated policy group** | **Policies:** Facial Coverings, Testing Policy, Vaccination Policy, Contact Tracing, Public Information Campaigns, Gathering Restrictions<br>**Effectiveness:** [-0.65]<br>**Efficiency:** [-0.0030]<br>**Total Cost:** \$208.8 Billion<br>**Effectiveness and Efficiency Index:** [0.42] |
| **All 13 Policy Group** | **Policies:** Facial Coverings, Testing Policy, Contact Tracing, Vaccination Policy, Internal Movement Restrictions, International Travel Controls, Public Information Campaigns, Public Event Cancellations, Gathering Restrictions, Closing Public Transportation, School Closures, Workplace Closures<br>**Effectiveness:** [-0.033]<br>**Efficiency:** [-0.0000061]<br>**Total Cost:** \$5381.8 Billion<br>**Effectiveness and Efficiency Index:** [0.019861] |

Through the above comparison analysis, as expected, a country implementing the most cost-efficient and effective policy group can save a tremendous amount of money while reducing the monthly case increase rate significantly more at the same time.

Comparing the results when Vaccinations were included and excluded, the research found that the vaccine policy can drastically improve the integrated efficiency and effectiveness index by around 58%. Facial Coverings and Contact Tracing are consistently cost-efficient and effective policies because they are part of the most efficient and effective policy group no matter if Vaccinations are included or not. In experiment 1, by adding Facial Coverings, it made the most efficient policy group about 42% more effective, with a similar cost efficiency (2.3% vs 2.7%).

## 5. Related Work

The work done by Wibbens, P. [21] utilizes Bayesian analysis on big data to evaluate the effectiveness of 11 widely implemented core COVID-19 policies in the U.S. It concluded that though these core policies reduced growth rates for new infections, they were still not enough to contain the virus. In order to bring the COVID-19 infection growth rate to near zero, higher-impact policies, like full workplace closures, would need to be implemented. The Bayesian approach is a method where experiment results are updated as more information is gathered. Since COVID-19 is an ongoing pandemic, Bayesian would be helpful in accounting for changes as time goes on. However, their research only focused on the effectiveness of policies in the United States, and does not consider the cost-efficiency of the policies. My research analyzed 13 common policies across 180 countries worldwide. Furthermore, my research focused more on the integrated cost-efficiency and effectiveness policy group and identified them through automatic machine-learning-enabled programs. Because very few countries can fight COVID-19 without considering a budget limitation, my research focusing on both cost-efficiency and effectiveness can provide more pragmatic and meaningful policy guidelines for countries, especially underdeveloped ones.

The work done by Doti, J. [4] analyzed the cost-benefits of statewide COVID-19 policy implementation in terms of lost jobs and real gross state product(RGSP). The data utilized was an Oxford COVID-19 policy stringency index - a numerical value to represent a state’s overall policy enforcement. It was concluded that despite policy interventions helping to reduce COVID-19 case rates, these policies caused millions to lose jobs and a $410 billion decline in RGSP. This work also estimated the national average cost per life to $1.1 million during this pandemic period. However, their work focused only on the U.S.’s COVID-19 circumstances and viewed policies as one large factor(a stringency index), while my work considered 180 countries around the world and analyzed 13 specific policies in both separate and integrated ways. Further, their research does not identify the most cost-efficient policies as my research did.

The work done by Arshed, N. et al. [1] utilized the Panel Random Coefficient Model to estimate the COVID-19 flattening curve and estimate the number of days it will take to reach the flattening point. It also evaluated the effectiveness of different COVID-19 policies around the world using Poisson regression, concluding that contact tracing, stay at home restrictions, and international movement restrictions are most effective in controlling spread and flattening the COVID-19 curve. My research identified the most effective policy group that includes Public Information Campaigns, Facial Coverings, Testing Policy, Contact Tracing, Vaccinations, Public Information Campaigns, Public Event Cancellations, and Stay at Home Requirements. A policy that their work deems effective but not my work is International Travel Controls. This difference could be a result of the different data and algorithms we used, as I used data across 180 countries from January 2020 to June 2021, but they used data from only January 2020 to May 2020; I also used Linear Regression while they used Poisson Regression. In addition, their research only focused on the effectiveness of policies and does not consider the cost-efficiency of the policies, on which my research focused on.

## 6. Conclusion

### 6.1 Summary

Based on the analysis of data on 13 common COVID 19 policies and daily case numbers across 180 countries in about 15 months, the research identified the most cost-efficient policy group, most effective policy group, and most cost-efficient and effective policy group through developing a policy cost model, defining policy cost-efficiency, policy effectiveness, and an integrated efficiency and effectiveness index, and running a developed best policies finder(a machine learning enabled computing program).

The research concluded that:

1. Based on the data on 12 common policies excluding Vaccinations,
  a. The most effective policy group(Facial Coverings, Testing Policy, Contact Tracing, Vaccinations, Public Event Cancellations, and Stay at Home Requirements)
    i. Effectiveness ∼ a 42% monthly case decrease rate
    ii. Cost-efficiency ∼ a 0.02% monthly case decrease rate per billion of dollars spent
    iii. Cost efficiency and effectiveness Index ∼ 0.25
  b. The most cost-efficient policy group (Facial Coverings, Testing Policy)
    i. Cost-efficiency ∼ a 1% monthly case decrease rate per billion of dollars spent
    ii. Effectiveness ∼ a 37% monthly case decrease rate
    iii. Cost efficiency and effectiveness Index ∼ 0.32
  c. The most cost-efficient and effective policy group (Facial Coverings, Testing Policy, Contact Tracing)
    i. Cost-efficiency ∼ a 0.9% monthly case decrease rate per billion of dollars spent
    ii. Effectiveness ∼ a 40% monthly case decrease rate
    iii. Cost efficiency and effectiveness Index ∼ 0.33
    iv. This policy group is 1474 times more cost-efficient, 11 times more effective, and costs around $5336.83 Billion less than implementing all 12 common COVID-19 policies as the U.S. and many other countries did before vaccinations were available.
2. Based on the data on all 13 common policies including Vaccinations,
  a. The hypothesis is incorrect. The all negative correlations policy group(Facial Coverings, Testing Policy, Vaccination Policy, Contact Tracing, Public Information Campaigns, Gathering Restrictions) is neither the most cost-efficient nor most effective. Its effectiveness(65%) is 76% of the most effective group(86%), and its efficiency(0.3%/$B) is only 11% of the most cost-efficient group(2.7%/$B). The all negative correlations policy group costs $189.17 billion more annually than the most cost-efficient policy group.
  b. The most effective policy group(Facial Coverings, Testing Policy, Contact Tracing, Vaccinations, Public Information Campaigns, Public Event Cancellations, and Stay at Home Requirements)
    i. Effectiveness ∼ a 86% monthly case decrease rate
    ii. Cost-efficiency ∼ a 0.03% monthly case decrease rate per billion of dollars spent
    iii. Cost efficiency and effectiveness Index ∼ 0.52
  c. The most cost-efficient policy group (Contact Tracing, Vaccination Policy)
    i. Cost-efficiency ∼ a 2.7% monthly case decrease rate per billion of dollars spent
    ii. Effectiveness ∼ a 52% monthly case decrease rate
    iii. Cost efficiency and effectiveness Index ∼ 0.58
  d. The most cost-efficient and effective policy group (Facial Coverings, Contact Tracing, and Vaccinations)
    i. Cost-efficiency ∼ a 2.2% monthly case decrease rate per billion of dollars spent
    ii. Effectiveness ∼ a 74% monthly case decrease rate
    iii. Cost efficiency and effectiveness Index ∼ 0.68
    iv. This policy group is 3835 times more cost-efficient, 21.5 times more effective, and costs around $5350 Billion less than implementing all 13 common COVID-19 policies as the U.S. and many other countries did.
3. In addition, Facial Covering and Contact Tracing are consistently efficient and effective policies because they are identified by the Best Policies Finder Program to be in the most cost-efficient and effective policy group, no matter whether the policy data includes or excludes Vaccinations.

### 6.2 Current Limitations

One main limitation of the research is that the cost model estimation is based on limited policy cost information available due to very little published data on COVID-19 policy costs. The current research estimated the policy cost through utilizing a wide variety of sources, and estimations with educated analyses. While some estimations may seem reasonable, like estimating the Facial Covering annual costs by multiplying the unit price of a mask by the total population of the U.S., other estimations may be rough since some policies’ costs are affected by many factors. This includes policies like Gathering Restrictions which can be affected by and correlated with Stay at Home Restrictions, Public Transportation Restrictions, International Travel Restrictions, and more. Another limitation of the research was that the linear regression prediction models’ R-Squared values varied from around 0.03 to 0.19, which is relatively low.

## 7. Future Work

To develop a more accurate cost model, it would be beneficial to continuously research and update the current policy cost model because there will be more data and research papers available on the cost-efficiency of the pandemic policies. Additionally, although linear regression is often applied in COVID-19 case prediction models development, it is not necessarily the best algorithm. To develop a better machine learning model for monthly case increase rate predictions with a higher R-Squared value on the same data set, future research may analyze, design, and test other possible machine learning algorithms.

## Data Availability

All data produced in the present study are available upon reasonable request to the authors.

